# An empirical assessment of influenza intensity thresholds obtained from the moving epidemic and WHO methods

**DOI:** 10.1101/2021.06.22.21259305

**Authors:** Johannes Bracher, Jonas M. Littek

**Affiliations:** Karlsruhe Institute of Technology (KIT), Chair of Statistics and Econometrics; Heidelberg Institute for Theoretical Studies (HITS), Computational Statistics Group

## Abstract

The moving epidemic method (MEM) and the WHO method are widely used to determine intensity levels for seasonal influenza. The two approaches are conceptually similar, but differ in two aspects. Firstly, the MEM involves a log transformation of incidence data, while the WHO method operates on the original scale. Secondly, the MEM uses more than one observation from each past season to compute intensity thresholds, fixing the total number to include. The WHO method uses only the highest value from each season. To assess the impact of these choices on thresholds we perform simulation studies which are based on re-sampling of ILI data from France, Spain, Switzerland and the US. When no transformation is applied, a rather large proportion of season peaks are classified as high or very high intensity. This can be mitigated by a logarithmic transformation. When fixing the total number of included past observations, thresholds increase the more seasons are available. When only few are available, there is a high chance of classifying new season peaks as high or very high intensity. We therefore suggest using one observation per season and a log transformation, i.e. a hybrid of the default settings of the MEM and WHO methods.

## 1 Introduction

Following the 2009 influenza H1N1 pandemic, the need for a rapid assessment tool for influenza intensity was recognized. The *Review Committee on the Functioning of the International Health Regulations and on Pandemic Influenza (H1N1)* recommended that member states perform yearly updates and evaluations of intensity thresholds (WHO, 2011, p.118). In the subsequent *WHO Pandemic Influenza Severity Assessment* (PISA) guideline (WHO, 2017) the so-called WHO method (WHO, 2014) and the moving epidemic method (MEM; Vega et al. 2013, 2015) were recommended to this end. The latter, which is also employed by the European Centre for Disease Prevention and Control (e.g., ECDC 2017), has been adopted by numerous national public health agencies (e.g., Dickson et al. 2020, Rakocevic et al. 2019, Redondo-Bravo et al. 2020, Vos et al. 2019); see Supplement C for an overview of recent applications. The statistical properties of the MEM and WHO methods, however, have not yet been studied in detail. We here address this aspect via simulation experiments based on re-sampling of historical influenza data. Our results indicate that it may be beneficial to use a hybrid of the default settings of these two methods.

## 2 Definition of the moving epidemic and WHO methods

We describe the computation of influenza/influenza-like illness (ILI) intensity thresholds, framing the MEM and WHO methods as two special cases of the same general approach. We assume that thresholds are based on weekly data (typically incidences) from *m* past seasons and applied to the (*m* + 1)-th season. Vega et al. (2015) recommend to use 5 ≤ *m* ≤ 10 seasons for the MEM to ensure a recent data basis. Computation of thresholds then proceeds as follows:

1. Select the *n* largest observations from each of the *m* past seasons to construct a set of reference observations.
2. Apply an (invertible) transformation to all selected observations.
3. Assume that the *m* × *n* transformed reference observations come from a normal distribution and compute estimates 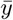, *s* of its mean and standard deviation.
4. Define intensity thresholds on the transformed scale as quantiles of the normal distribution 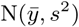. A common choice for both methods is
  - the 40th percentile 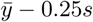 as the threshold between low and medium intensity;
  - the 90th percentile 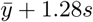 as the threshold between medium and high intensity;
  - the 97.5th percentile 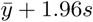 as the threshold between high and very high intensity.
5. Transform thresholds back to the original scale (e.g. using the exponential function if a log transformation was used).

The MEM and WHO methods are special cases of this procedure:

- In the WHO method, *n* = 1 observation per season is used and by default no transformation is applied. If peak incidences vary strongly across seasons, a log transformation is recommended (WHO, 2017).
- In the MEM method, the default transformation is the natural logarithm, while the number of included observations per season is set to *n* = 30*/m*, rounded to the nearest integer. The total number of historical observations is thus kept approximately fixed.

The rationale behind the percentile-based approach is that “about 50–60% of the season peaks should be above the moderate threshold, ±10% above the high threshold and ±2.5% above the extraordinary threshold” (WHO, 2017, p.10).

The implementation of the moving epidemic method in the R package `mem` (Lozano, 2020) permits the user to choose *n, m*, and *f* (`i.type.intensity = 5` for no transformation, `i.type.intensity = 6` for the log transformation). The term *moving epidemic method* could thus also be used as an umbrella term for the general procedure described above. We here use it in a more narrow sense for the specification from Vega et al. (2015), reflected in the default settings of the `mem` package.

In previous works (WHO 2014; Vega et al. 2015), the above thresholds have been described as upper ends of one-sided confidence intervals for the arithmetic (WHO method) or geometric mean (MEM) of the reference observations. This, however, is imprecise terminology as in the computations the standard deviation *s* rather than the standard error 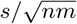 is used (see documentation of the `mem` package and WHO 2014, p.69). We note that the use of actual confidence intervals is also possible in the `mem` package (`i.type.intensity = 1` for the geometric, `i.type.intensity = 2` for the arithmetic mean). However, thresholds for all levels will then converge to the arithmetic or geometric mean if sufficient historical data are available. As illustrated in Supplement B, this is not a desirable behaviour.

## 3 Simulation study

### 3.1 Goal

We aim to assess how thresholds depend on the employed transformation, the number of observations *n* used per season and the number *m* of seasons included. In particular, the following aspects are of interest:

- The application of a logarithmic transformation is expected to lead to higher thresholds for the extreme categories and thus a lower proportion of seasons peaks classified as high or very high.
- In the MEM, the reference set also contains past observations which are not actually peak observations. This may introduce a downward bias in thresholds, especially if due to a small number *m* of available seasons *n* is large.

The latter aspect is intuitive, but also supported by formal statistical considerations detailed in Supplement A. These imply that thresholds are unbiased for *n* = 1 in the sense that, as intended, they will be exceeded by around 60%/10%/2.5% of the season peaks in the long run (if some auxiliary assumptions are fulfilled). When *n >* 1, thresholds will tend to be lower and exceeded more frequently.

### 3.2 Simulation setup

We compare four versions of the general approach described in Section 2:

a. No transformation, *n* = 1. This corresponds to the WHO method.
b. No transformation, *n* = 30*/m*.
c. Logarithmic transformation, *n* = 1.
d. Logarithmic transformation, *n* = 30*/m*. This corresponds to the MEM approach.

In order to closely mimic the seasonal patterns of influenza, we re-sample historical surveillance data rather than generating fully synthetic data. Assume *M* seasons of historical data on a measure of influenza activity are available. We then repeat the following steps 500 times:

- Sample a sequence of 15 seasons from the *M* available seasons. This is done with equal probability for each season and *with replacement*, meaning that the same season can appear more than once. This approach is called the *seasonal block bootstrap* (Politis, 2001).
- For each value *m* = 5, …, 15:
  – Restrict the generated sequence to the first *m* seasons.
  – Apply approaches (a)–(d) to compute thresholds for medium (40th percentile), high (90th percentile) and very high intensity (97.5th percentile).
  – Compute which fraction of all *M* available historical season peaks would be classified as low, moderate, high and very high.

The range *m* = 5, …, 15 is motivated by the range of values found in real-world applications, see overview in Supplement C. All analyses were performed using the R language for statistical computing (R Core Team, 2020) and the package `mem` (Lozano, 2020).

### 3.3 Data

We use publicly available influenza surveillance data from four countries. Data on the weekly incidence of influenza-like illness in France, 1985–2019, were obtained from Réseau Sentinelles (INSERM/Sorbonne Université, https://www.sentiweb.fr, Flahault et al. 2006). Data on weekly confirmed influenza cases in Spain, 1998–2019, were extracted from graphs shown in the *Informe Anual* of the *Sistema de la vigilancia de gripe en España* (https://vgripe.isciii.es/; Sistema de Vigilancia de Gripe en España 2019). Weekly ILI counts from Switzerland, 2000–2016, collected by the Swiss Federal Office of Public Health are available in the R package `HIDDA.forecasting` (Held and Meyer, 2019). Weekly weighted ILI (wILI) data at the US national level from CDC FluView (Charbonneau and James, 2019), 1998–2017, were obtained via the CDC FluSight influenza forecasting platform (https://github.com/FluSightNetwork/cdc-flusight-ensemble/). These wILI values correspond to the fraction of general practitioner visits due to influenza-like symptoms. All four time series are displayed in Figure 1, with the pandemic 2009/2010 season removed. We also show boxplots of the first through sixth largest observation per season. Not surprisingly, values on average get smaller for increasing ranks. Interestingly, they also get less dispersed, meaning that variability among e.g. the sixth largest observations per season is smaller than among the peak values.

**Figure 1:**
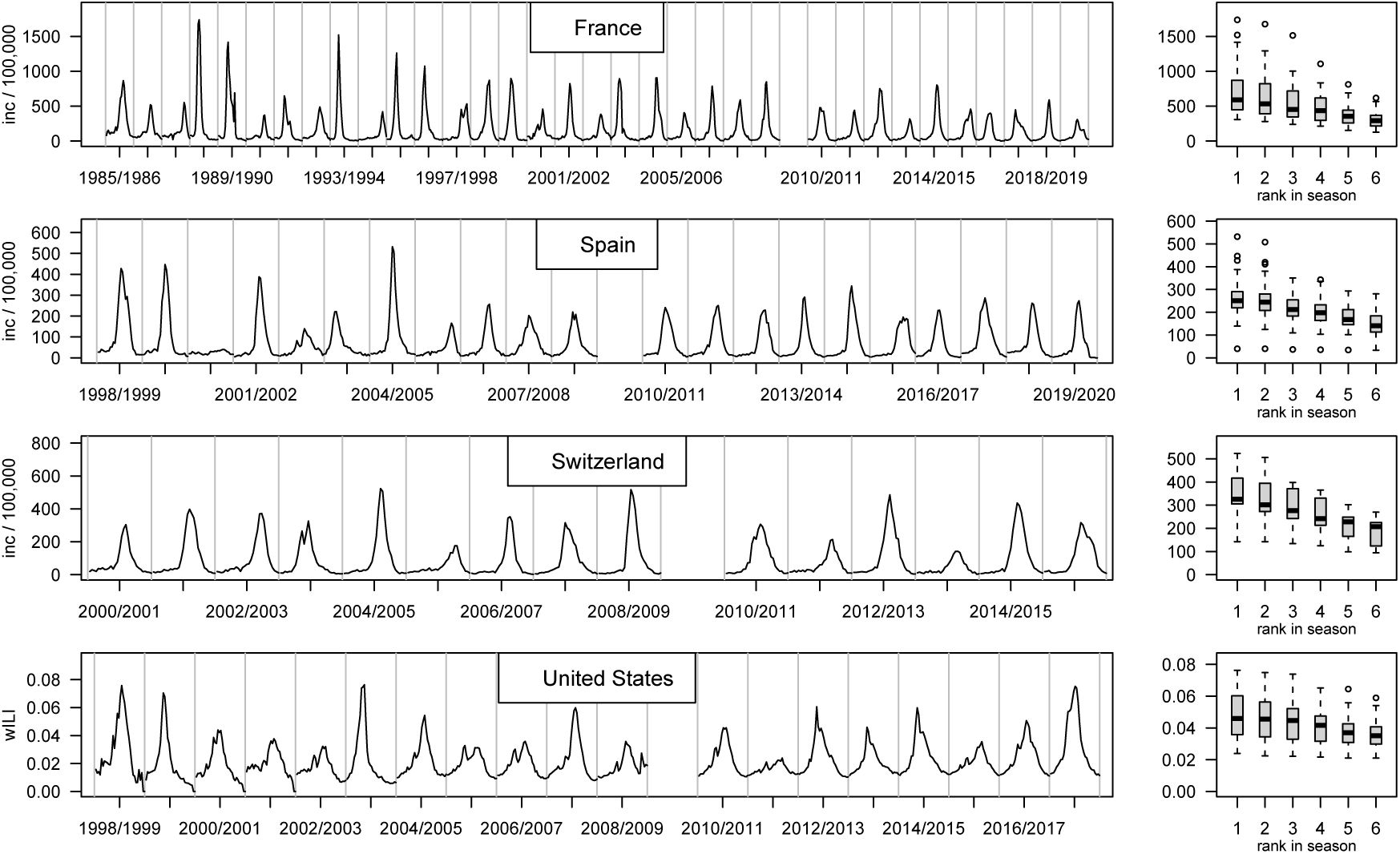
Time series of influenza activity measures in four countries: Weekly ILI cases per 100,000 population in France, 1985–2019; weekly number of confirmed influenza cases per 100,000 population in Spain, 1998–2019; weekly ILI cases per 100,000 in Switzerland, 2000–2016; weekly wILI values in the United States, 1998–2017. Off-season weeks are omitted in the plot, with grey lines delimiting the different seasons. The right column shows boxplots of the largest value per season, second largest etc.

### 3.4 Results

Results from our simulation study are shown in Figures 2 (France, Spain) and 3 (Switzerland and US). In the first and third column of each figure we also show an analytical approximation of the expected thresholds, which agrees very well with the simulation results; see Supplement A for details. In all four countries, using a log transformation leads to increased thresholds, in particular for the high and very high levels. Especially in France and Spain, thresholds obtained without the log transformation are too low, as can be seen from the large proportion of season peaks classified as very high (around 10% when using *n* = 1). As expected, when letting the number of observations used per season depend on the number of available seasons via *n* = 30*/m*, average thresholds increase in *m*. As a particularly striking example, the average threshold for high intensity in France (when using a log transformation) increases from 834 for *m* = 5, *n* = 6 to 1011 for *m* = 10, *n* = 3 and 1080 for *m* = 15, *n* = 2. For *n* = 1, in which case thresholds can be interpreted as unbiased (Supplement A), the average is 1147. Including historical observations which are not from actual peak weeks thus leads to a considerable lowering of alarm thresholds and will increase the number of alerts for high and very high influenza activity. This is not surprising given the pronounced differences between the distribution of season peak values and e.g. fifth largest observation per season, see right column of Figure 1. In the US and France, on average one in three season peaks is classified as high and one in seven as very high intensity when applying the MEM with log transformation and *m* = 5, *n* = 6. This is rather far from the intended exceedance probabilities of 10% and 2.5%.

**Figure 2:**
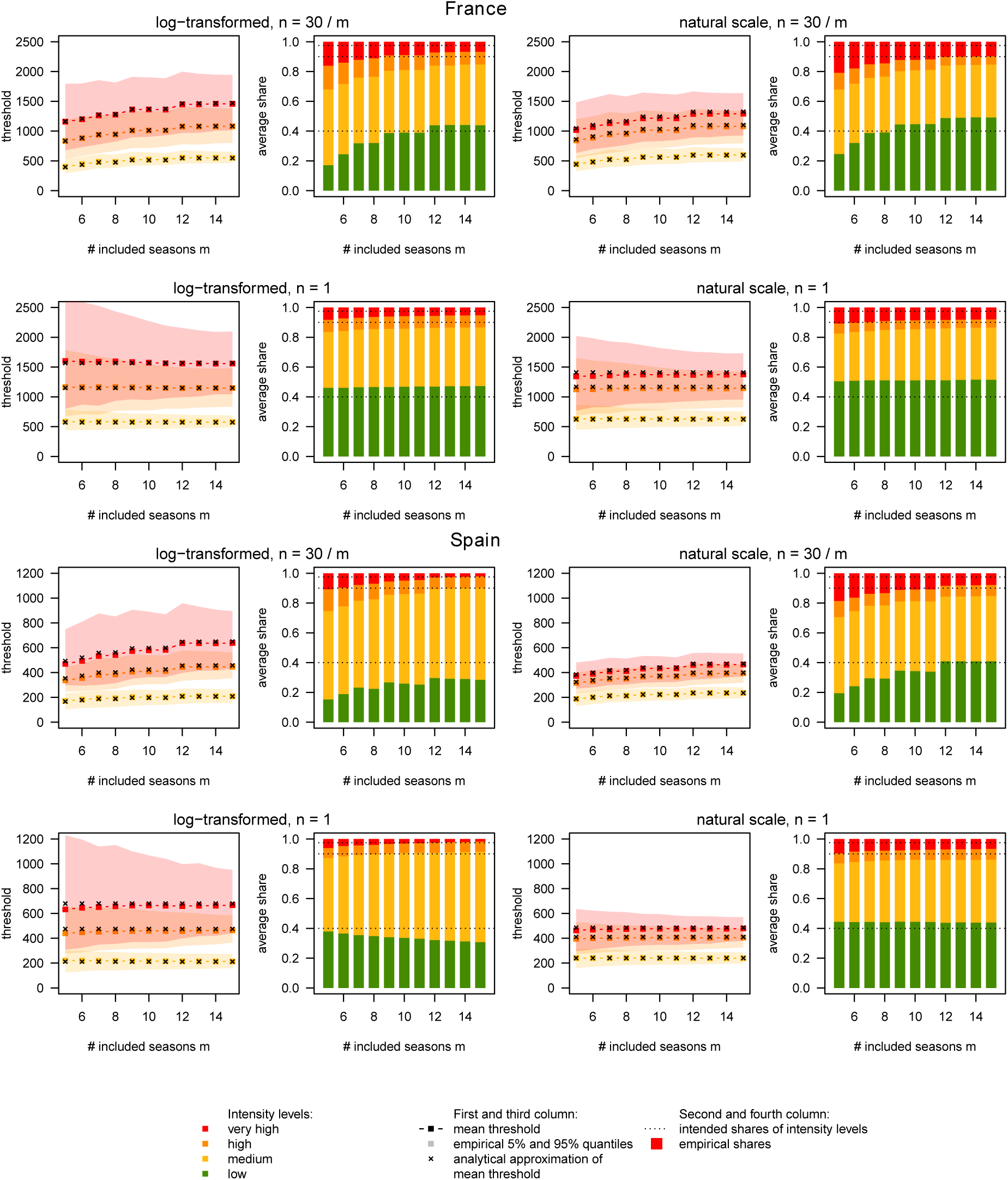
Results of simulation study, France and Spain. First and third column: mean intensity thresholds along with bands delimited by the empirical 5% and 95% quantiles. Analytical approximations of the mean threshold values are marked as small black crosses. Second and fourth columns: resulting average shares of season peaks classified as low, medium, high and very high intensity.

When always using *n* = 1, the average thresholds and shares of the different categories are more well-behaved also for small *m*. This holds especially when applying a log transformation, even though certain mismatches with the nominal exceedance probabilities remain. Also, there is considerable variability in the estimated thresholds (shaded areas in Figures 2 and 3). These difficulties, however, are inherent in the problem of estimating a 90% or 97.5% quantile based on 5–10 observations.

**Figure 3:**
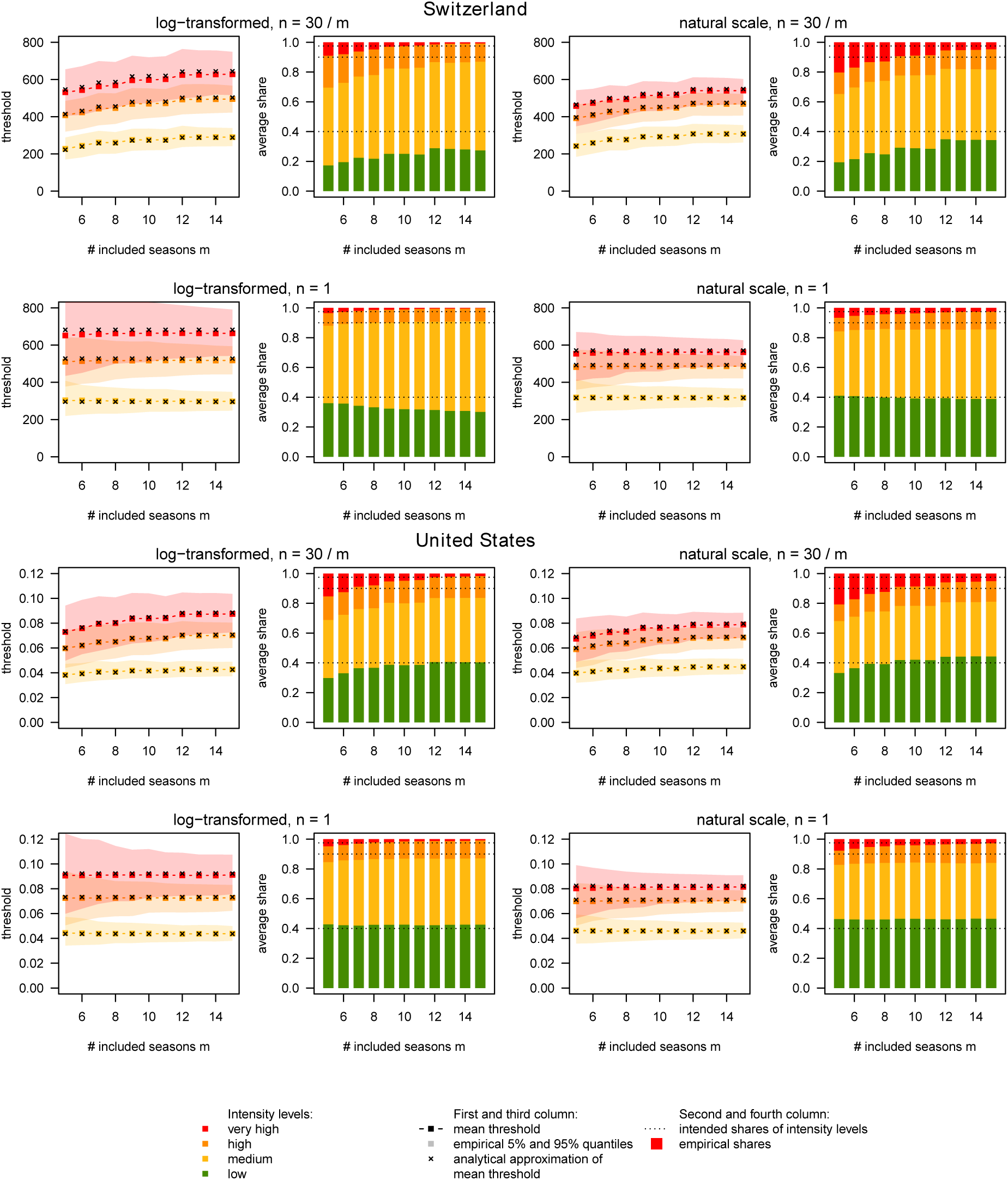
Results of simulation study, Switzerland and United States. First and third column: mean intensity thresholds along with bands delimited by the empirical 5% and 95% quantiles. Analytical approximations of the mean threshold values are marked as small black crosses. Second and fourth columns: resulting average shares of season peaks classified as low, medium, high and very high intensity.

## 4 Discussion

We provided a statistical assessment of implementation choices in a widely used framework for the computation of influenza intensity thresholds. We found that applying a log transformation leads to better behaved thresholds and closer to nominal exceedance rates. Concerning the question of how many observations per historical season should be included to compute thresholds, we found that the common choice *n* = 30*/m* results in too low thresholds when few historical seasons are available. We therefore recommend adopting *n* = 1 irrespective of the number of available historical seasons. This is possible in the `R` package `mem` by setting `i.n.max = 1` when the function `memmodel` is called.

We think that a simple and interpretable tool with a well-documented and open source software implementation like `mem` is a valuable tool in practice. The use of a standard approach at the European level will improve comparability of results, facilitating the evaluation and refinement of the tool. With this work we hope to contribute a statistical perspective on this topic, complementing public health practitioners’ experience from applied analyses.

## Data Availability

Materials to reproduce the presented results are available at https://github.com/jbracher/mem.

## Data and code

Materials to reproduce the presented results are available at https://github.com/jbracher/mem.

## Ethics statement

No ethics approval was necessary as this study uses exclusively publicly available data.

## Acknowledgements

We would like to thank Laura Werlen and Daniel Wolffram for helpful feedback. Johannes Bracher was supported by the Helmholtz Foundation via the SIMCARD Information and Data Science Pilot Project.

## Supplementary Materials

This supplementary material is hosted by Eurosurveillance as supporting information alongside the article *An empirical assessment of influenza intensity thresholds obtained from the moving epidemic and WHO methods*, on behalf of the authors, who remain responsible for the accuracy and appropriateness of the content. The same standards for ethics, copyright, attributions and permissions as for the article apply. Supplements are not edited by Eurosurveillance and the journal is not responsible for the maintenance of any links or email addresses provided therein.

### A Formal statistical analysis

#### A.1 Re-stating the definitions

We start by re-stating the definitions of the MEM and WHO method in a slighty more formal way and introducing relevant notation. We assume again that thresholds are based on data from *m* past seasons and applied to the (*m* + 1)-th season. Thresholds for an intensity measure *X* are then obtained as follows.

1. Within each historical season *j* = 1, …, *m* order all observations in decreasing order, denoting the *i*-th largest observation from season *j* by 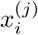.
2. Select the *n* largest observations from each of the *m* past seasons to construct a reference set 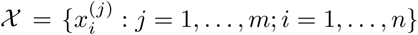.
3. Apply an (invertible) transformation 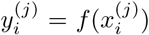 to all members of the reference set 𝒳 to obtain a reference set 𝒴 of transformed historical observations.
4. Assume that the transformed values in 𝒴 come from a normal distribution and compute estimates 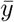, *s* of its mean and standard deviation.
5. Define intensity thresholds on the transformed scale as quantiles of the normal distribution 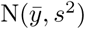, i.e. compute

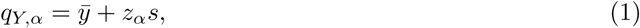

where *z*_*α*_ is the *α* quantile of the standard normal distribution. A common choice for both methods is
  - the 40th percentile 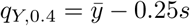 as the threshold between low and medium intensity;
  - the 90th percentile 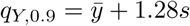 as the threshold between medium and high intensity;
  - the 97.5th percentile 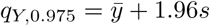 as the threshold between high and very high intensity.
6. Obtain thresholds on the original scale by applying the inverse transformation, i.e. setting *q*_0.4_ = *f* ^*−*1^(*q*_*Y*,0.4_), *q*_0.9_ = *f* ^*−*1^(*q*_*Y*,0.9_), *q*_0.975_ = *f* ^*−*1^(*q*_*Y*,0.975_).

The MEM and WHO methods are special cases of this procedure with the following specifications:

- In the WHO method, *n* = 1 observation per season is used irrespective of the number *m* of historical seasons. The standard procedure is to apply no transformation, i.e. set *f* to the identity function.
- In the MEM method, the default choice for *f* is the natural logarithm, while the number of observations per season is set to *n* = 30*/m*. (The exact specification used in the R package, `mem` is *n* = max(1, ⌊ 30*/m* ⌋), where ⌊ ⌋ denotes rounding to the nearest integer.) The total number of historical observations is thus kept approximately fixed.

#### A.2 Results

We assume that the different historical seasons are independent realizations of the same random process, which is certainly not strictly true, but should hold in good approximation. Denote by **Y**^(*j*)^ the random vector of the *n* largest transformed incidences from season *j* in decreasing order, i.e. 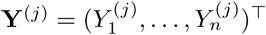 with 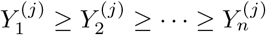. The mean and covariance matrix of **Y**^(*j*)^ are denoted by

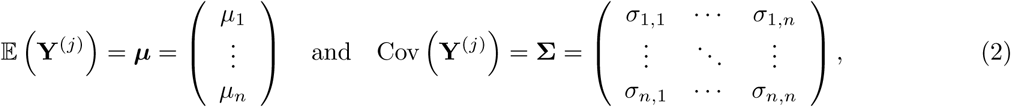

respectively, where to make notation more intuitive we also write 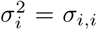. It can then be shown that the expectations of 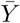 and *S*^2^ when using *m* seasons and *n* observations from each season are given by

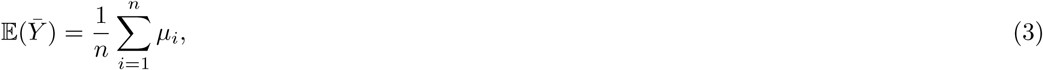

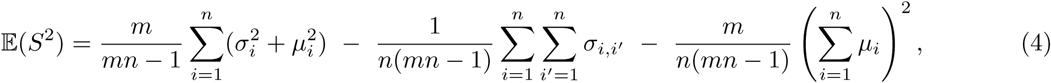

respectively. The derivation is provided in the next subsection. For reasons detailed there, if the transformation function *f* is the identity or the natural logarithm,

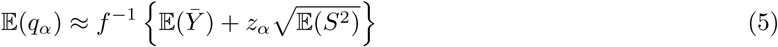

usually holds in good approximation in our applied setting. It can be shown that for *n* = 1, we have

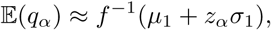

usually in good approximation. This is a desirable property. Loosely speaking, if in addition the 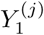 come from a normal distribution, the nominal exceedance probabilities (60%/10%/2.5%) for the different thresholds would in the long run be achieved. If a different value *n >* 1 is chosen, this will generally no longer be the case. Equations (3)–(5) tell us by how much *q*_*α*_ can be expected to differ from the intended value *f* ^*−*1^(*µ*_1_ + *z*_*α*_*σ*_1_). By the definition of the *µ*_*i*_ (with *µ*_*i*_ the expectation of the *i*-th largest observation in a given season), 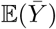 decreases in *n*. In most (possibly all) settings this also translates to the *q*_*α*_. As a consequence, when choosing *n >* 1, one must expect to classify a larger number of season peaks as high or very high intensity. When choosing *n* = 30*/m* as suggested for the MEM, thresholds will then tend to increase the more years of data are used to compute thresholds.

#### A.3 Derivation

We start by addressing the expectations of empirical mean 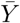 and variance *S*^2^ of the reference observations, where

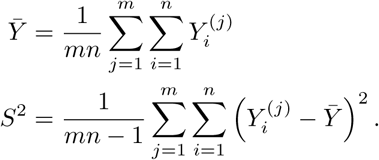

It is straightforward to see that

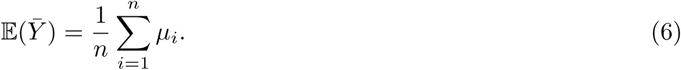

For the variance *S*^2^, we first note that it can be re-written as

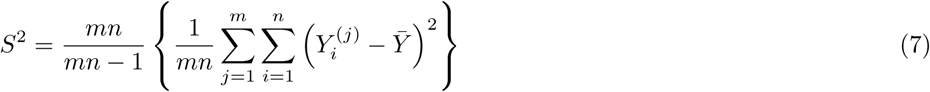

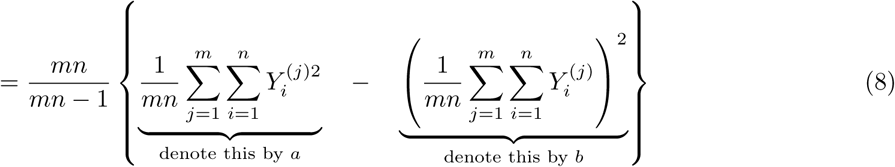

We consider the two terms *a* and *b* separately, starting by

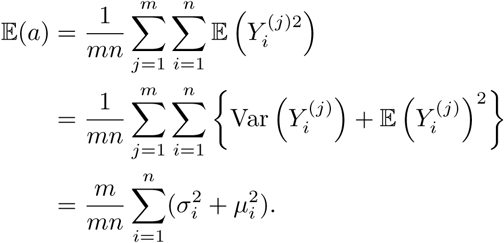

Then we note that

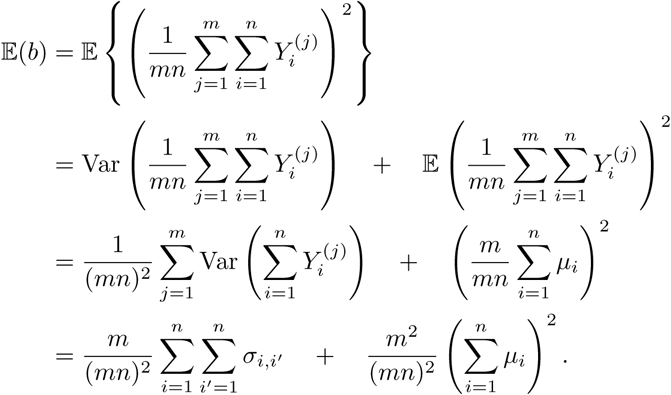

Plugging these results back into equation (8) we obtain

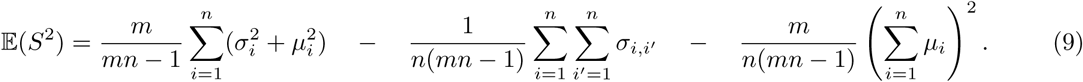

It is straightforward to see that for *n* = 1 the expressions (3) and (9) simplify to

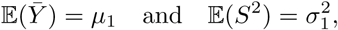

as given in equation (4).

There is no general way of computing the expectation 𝔼 (*S*) from the respective standard deviation, but unless the true distribution of *S*^2^ has strong excess curtosis,

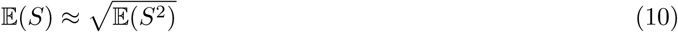

is a reasonable approximation. We can then plug equations (3) and (10) into the formulae for the thresholds *q*_*Y,α*_ on the transformed scale and obtain

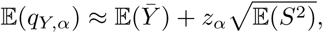

where *z*_*α*_ is the *α* quantile of the standard normal distribution (with *α* ∈ {0.4, 0.9, 0.975}).

If *f* was set to the natural logarithm, the question remains how to obtain statements concerning thresholds *q*_*α*_ on the original scale. Approximation via a second-order Taylor expansion yields

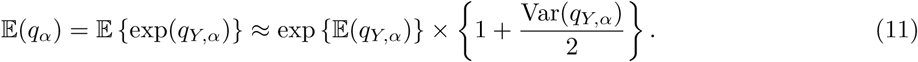

Empirically, after transformation to the log scale, the variance of the reference observations is low in our applied setting. The resulting variances of *q*_*Y,α*_ are then quite small and do not play an important role in equation (11). We can thus use the even simpler approximation

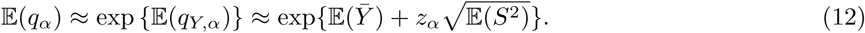

As can be seen from Figures 2 and 3 from the main manuscript, this approximation works very well in praxis. To compute the values indicated by the small black crosses, we just plugged the empirical mean vectors and covariance matrices of the **y**^(*j*)^ in the respective data sets into equations (3)–(5).

### B Intensity thresholds based on confidence intervals

Previous works have referred to the thresholds discussed in our manuscript as upper ends of one-sided confidence intervals associated with the arithmetic (WHO) or geometric mean (MEM) of the reference observations (WHO 2014; Vega et al. 2015). This, however, is an imprecise use of terminology as the thresholds are computed using the standard deviation *s*. Confidence intervals, on the other hand, would be computed using the standard error 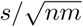. A more precise term would thus be “prediction interval”, which implies that the interval refers to one future realization rather than a theoretical mean.

These terminology questions aside, we note that the `mem` R package also offers threshold computation based on actual confidence intervals (`i.type.intensity = 1` for the geometric, `i.type.intensity = 2` for the arithmetic mean). On the transformed scale these are computed as

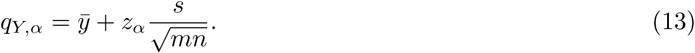

These thresholds show somewhat peculiar behaviour. We illustrate this by repeating the simulation study the main manuscript for France with the respective settings. The result is shown in Figure S1.

**Figure S1:**
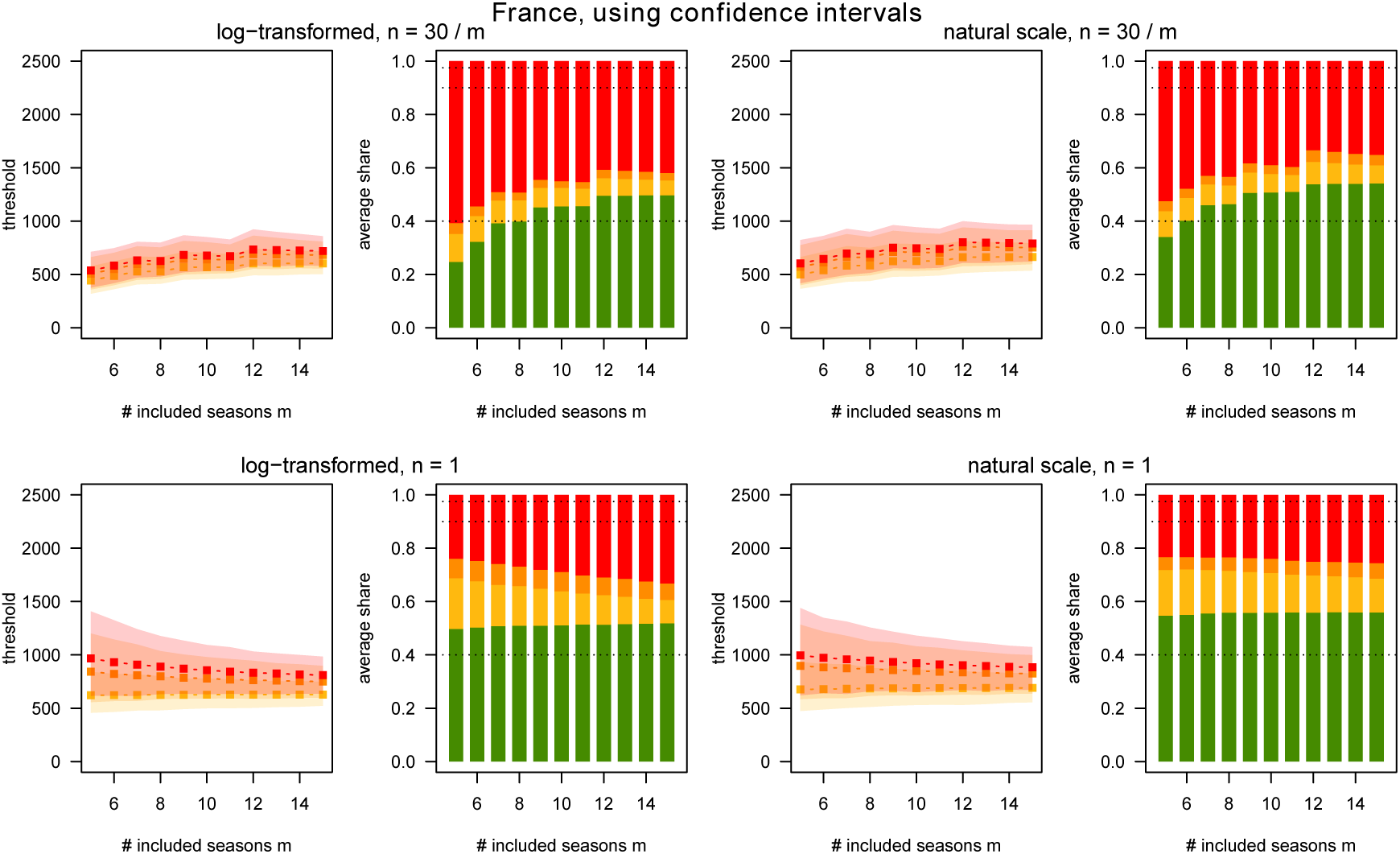
Simulation results for France as in Figure 2 of the main manuscript, but using confidence intervals based on standard errors rather than prediction intervals based on standard deviations.

Here, two different mechanisms are at work. If one chooses *n* = 1 per season irrespective of the number *m* of available seasons, the number of included observations *mn* increases in *m*. With growing *m* this leads to confidence intervals which get narrower and a funnel-like pattern in the different thresholds (bottom row of Figure S1). If one chooses *n* = 30*/m*, the number *mn* will remain constant (or approximately, as some rounding is necessary). The funnel-like shape is thus not observed and an increasing pattern like in our main analyses emerges instead. In both cases, there is an excessive number of seasons classified as very high intensity. This reflects the fact that the extreme thresholds necessarily get smaller when using the standard error rather than standard deviation in equation (13). We thus conclude that these variants of the moving epidemic method are not advisable in practice.

### C An overview of recent applications

To get a better understanding of the different settings in which the MEM and WHO methods are applied in practice we performed a literature search of articles published in English and citing the papers Vega et al. (2015), WHO (2014) and WHO (2017) until December 2020 (identified via *CrossRef* and *Google Scholar*). The results are summarized in Table S1.

**Table S1:** Applications of the MEM and WHO method to determine intensity thresholds for respiratory diseases. We did not include works where only baseline thresholds are computed. The number of seasons included to compute thresholds is denoted by *m*, the number of observations used per season by *n*. The “Percentiles” column indicates which percentiles were used for the medium, high and very high thresholds, with “?” indicating that no explicit information was found. Abbreviations: SARI = severe acute respiratory infection; ILI = influenza-like illness; RSV = respiratory syncytial virus.

| (a) Moving epidemic method |  |  |  |  |  |  |
| --- | --- | --- | --- | --- | --- | --- |
| Region | Disease | Years covered | $m$ | $n$ | Percentiles | Authors |
| Australia | ILI/influenza | 2012–2017 | 5 | 6 | 40, 90, 99 | Vette et al. (2018) |
| Australia, Chile, New Zealand, South Africa | ILI/SARI | 2013–2019 | 6 | 5 | 40, 90, 97.5 | Sullivan et al. (2019) |
| Catalonia | ILI | 2010–2016 | 5 | 6 | ? | Basile et al. (2018) |
| Catalonia | ILI | 2005–2018 | 12 | 3 | ? | Basile et al. (2019) |
| Catalonia | ILI/influenza | 2010–2017 | 7 | 4 | ? | Torner et al. (2019) |
| Egypt | SARI/ILI | 2010–2017 | 6 | 5 | 40, 90, 97.5 | AbdElGawad et al. (2020) |
| Egypt | SARI | 2013–2015 | 3 | 10 | ? | Elhakim et al. (2019) |
| England | ILI | 2010–2016 | 6 | 5 | ? | Wagner et al. (2018) |
| Finland | influenza | 2011–2016 | 5 | 6 | ? | Pesälä et al. (2019) |
| Montenegro | ILI | 2010–2018 | 7 | 4 | 40, 90, 97.5 | Rakocevic et al. (2019) |
| Morocco | ILI | 2005–2017 | 11 | 3 | 40, 90, 97.5 | Rguig et al. (2020) |
| Netherlands | RSV | 2005–2017 | 12 | 3 | 40, 90, 97.5 | Vos et al. (2019) |
| Norway | influenza | 2006–2015 | 9 | 3 | ? | Benedetti et al. (2019) |
| Pakistan | ILI, SARI | 2008–2017 | 10 | 3 | 40, 90, 97.5 | Nisar et al. (2020) |
| Portugal | ILI | 2012–2017 | 5 | 6 | 40, 90, 97.5 | Páscoa et al. (2018) |
| Scotland | influenza | 2010–2018 | 7 | 4 | ? | Murray et al. (2018) |
| Scotland | influenza | 2010–2019 | 7–8 | 4 | 40, 90, 97.5 | Dickson et al. (2020) |
| Slovenia | RSV | 2008–2018 | 10 | 3 | 40, 90, 97.5 | Grilc et al. (2021) |
| Spain (17 regions) | ILI | 2003–2015 | 4–10 | 3–8 | 40, 90, 97.5 | Bangert et al. (2017) |
| Spain | ILI | 2001–2018 | 16 | 2 | 40, 90, 97.5 | Redondo-Bravo et al. (2020) |
| Tunisia | ILI | 2009–2018 | 9 | 3 | 50, 90, 95 | Bouguerra et al. (2020) |
| United Kingdom | ILI | 2000–2013 | 10 | 3 | 40, 90, 97.5 | Green et al. (2015) |
| United Kingdom | ILI/RSV | 2011–2018 | 4–6 | 5–8 | 40, 90, 97.5 | Harcourt et al. (2019) |
| USA | ILI/influenza | 2003–2015 | 11 | 3 | 50, 90, 98 | Biggerstaff et al. (2017) |
| USA | ILI | 2010–2015 | 5 | 6 | 50, 90, 98 | Dahlgren et al. (2018) |
| USA | influenza | 2010–2016 | 6 | 5 | 50, 90, 98 | Dahlgren et al. (2019) |
| (b) WHO method |  |  |  |  |  |  |
| Region | Disease | years covered | $m$ | $n$ | percentiles | authors |
| Cambodia | ILI | 2009–2015 | 7 | 1 | mean, 90, 95 | Ly et al. (2017) |
| Morocco | ILI | 2005–2017 | 11 | 1 | 40, 90, 97.5 | Rguig et al. (2020) |
| Philippines | ILI | 2006–2012 | 7 | 1 | 90 | Lucero et al. (2016) |
| Victoria/Australia | ILI | 2002–2011 | 6–10 | 1 | 90, 95 | Tay et al. (2013) |

As can be seen from the large number of entries from the years 2019 and 2020, the MEM has quickly become a standard approach in the determination of intensity thresholds for influenza and other respiratory diseases. Indeed, the contributions come from numerous countries and in many cases have been co-authored by representatives of national or regional public health agencies. In most analyses, the suggested threshold levels at the 40th, 90th and 97.5th percentile as described in Section A.1 are used. Variability with respect to the number *m* of historical seasons included is considerable, with a range from 3 to 16 seasons. Consequently, the number *n* of observations included per season ranges from two to ten (none of the above papers indicated a modification of the default setting *n* = 30*/m* of the moving epidemic method). We only found three published applications of the WHO method, one of them providing a comparison to the thresholds from the MEM method (Rguig et al., 2020).

## References

Charbonneau, D. H. and James, L. N. (2019). FluView and FluNet: Tools for influenza activity and surveillance. Medical Reference Services Quarterly, 38(4):358–368. PMID: 31687905.

Dickson, E. M., Marques, D. F., Currie, S., Little, A., Mangin, K., Coyne, M., Reynolds, A., McMenamin, J., and Yirrell, D. (2020). The experience of point-of-care testing for influenza in Scotland in 2017/18 and 2018/19 – no gain without pain. Eurosurveillance, 25(44).

ECDC (2017). Risk assessment for seasonal influenza, EU/EEA, 2017–2018. Available online at https://www.ecdc.europa.eu/sites/default/files/documents/RRA%20seasonal%20influenza%20EU%20EEA%202017-2018-rev_0.pdf. Last accessed 27 December 2020.

Flahault, A., Blanchon, T., Dorléans, Y., Toubiana, L., Vibert, J. F., and Valleron, A. J. (2006). Virtual surveillance of communicable diseases: a 20-year experience in France. Statistical Methods in Medical Research, 15(5):413–421. PMID: 17089946.

Held, L. and Meyer, S. (2019). Forecasting based on surveillance data. In Held, L., Hens, N., O’Neill, P. D., and Wallinga, J., editors, Handbook of Infectious Disease Data Analysis. Chapman and Hall/CRC.

Lozano, J. (2020). mem: The moving epidemic method. R package, version 2.16 available via CRAN, https://cran.r-project.org/web/packages/mem/index.html.

Politis, D. (2001). Resampling time series with seasonal components. Frontiers in Data Mining and Bioinformatics: Proceedings of the 33rd Symposium on the Interface of Computing Science and Statistics: Orange County, California, June 13-17, page 619–621.

R Core Team (2020). R: A Language and Environment for Statistical Computing. R Foundation for Statistical Computing, Vienna, Austria.

Rakocevic, B., Grgurevic, A., Trajkovic, G., Mugosa, B., Grujicic, S. S., Medenica, S., Bojovic, O., Alonso, J. E. L., and Vega, T. (2019). Influenza surveillance: determining the epidemic threshold for influenza by using the Moving Epidemic Method (MEM), Montenegro, 2010/11 to 2017/18 influenza seasons. Eurosurveillance, 24(12):1800042.

Redondo-Bravo, L., Delgado-Sanz, C., Oliva, J., Vega, T., Lozano, J., Larrauri, A., and the Spanish Influenza Sentinel Surveillance System (2020). Transmissibility of influenza during the 21st-century epidemics, Spain, influenza seasons 2001/02 to 2017/18. Eurosurveillance, 25(21).

Sistema de Vigilancia de Gripe en España (2019). Guía de procedimientos para la vigilancia de gripe en España. Available online at https://www.isciii.es/QueHacemos/Servicios/VigilanciaSaludPublicaRENAVE/EnfermedadesTransmisibles/Documents/GRIPE/GUIAS/Gu%C3%ADa%20de%20procedimientos%20para%20la%20vigilancia%20de%20gripe%20en%20Espa%C3%B1a_marzo%202019.pdf. Last accessed 13 January 2020.

Vega, T., Lozano, J. E., Meerhoff, T., Snacken, R., Beauté, J., Jorgensen, P., Ortiz de Lejarazu, R., Domegan, L., Mossong, J., Nielsen, J., Born, R., Larrauri, A., and Brown, C. (2015). Influenza surveillance in Europe: comparing intensity levels calculated using the Moving Epidemic Method. Influenza and Other Respiratory Viruses, 9(5):234–246.

Vega, T., Lozano, J. E., Meerhoff, T., Snacken, R., Mott, J., Ortiz de Lejarazu, R., and Nunes, B. (2013). Influenza surveillance in Europe: establishing epidemic thresholds by the Moving Epidemic Method. Influenza and Other Respiratory Viruses, 7(4):546–558.

Vos, L. M., Teirlinck, A. C., Lozano, J. E., Vega, T., Donker, G. A., Hoepelman, A. I., Bont, L. J., Oosterheert, J. J., and Meijer, A. (2019). Use of the moving epidemic method (MEM) to assess national surveillance data for respiratory syncytial virus (RSV) in the Netherlands, 2005 to 2017. Eurosurveillance, 24(20):1800469.

WHO (2011). Strengthening response to pandemics and other public-health emergencies: Report of the review committee on the functioning of the international health regulations (2005) and on pandemic influenza (H1N1) 2009. Available online at https://www.who.int/ihr/publications/RC_report/en/. Last accessed 26 December 2020.

WHO (2014). WHO global epidemiological surveillance standards for influenza. Available online at https://www.who.int/influenza/resources/documents/influenza_surveillance_manual/en/ (last accessed 27 December 2020).

WHO (2017). Pandemic influenza severity assessment (PISA). Available online at https://apps.who.int/iris/handle/10665/259392. Last accessed 27 December 2021.

## References

AbdElGawad, B., Vega, T., El Houssinie, M., Mohsen, A., Fahim, M., Abu ElSood, H., Jabbour, J., Eid, A., and Refaey, S. (2020). Evaluating tools to define influenza baseline and threshold values using surveillance data, Egypt, season 2016/17. Journal of Infection and Public Health, 13(3):430 – 437.

Bangert, M., Gil, H., Oliva, J., Delgado, C., Vega, T., De Mateo, S., and Larrauri, A. (2017). Pilot study to harmonize the reported influenza intensity levels within the Spanish influenza sentinel surveillance system (SISSS) using the Moving Epidemic Method (MEM). Epidemiology and Infection, 145(4):715–722.

Basile, L., Oviedo de la Fuente, M., Torner, N., Martínez, A., and Jané, M. (2018). Real-time predictive seasonal influenza model in Catalonia, Spain. PLOS ONE, 13(3):1–15.

Basile, L., Torner, N., Martínez, A., Mosquera, M., Marcos, M., and Jane, M. (2019). Seasonal influenza surveillance: Observational study on the 2017–2018 season with predominant B influenza virus circulation. Vacunas, 20(2):53 – 59.

Benedetti, G., White, R. A., Pasquale, H. A., Stassijns, J., van den Boogaard, W., Owiti, P., and Van den Bergh, R. (2019). Identifying exceptional malaria occurrences in the absence of historical data in South Sudan: a method validation. Public Health Action.

Biggerstaff, M., Kniss, K., Jernigan, D. B., Brammer, L., Bresee, J., Garg, S., Burns, E., and Reed, C. (2017). Systematic assessment of multiple routine and near real-time indicators to classify the severity of influenza seasons and pandemics in the United States, 2003–2004 through 2015–2016. American Journal of Epidemiology, 187(5):1040–1050.

Bouguerra, H., Boutouria, E., Zorraga, M., Cherif, A., Yazidi, R., Abdeddaiem, N., Maazaoui, L., ElMoussi, A., Abid, S., Amine, S., Bouabid, L., Bougatef, S., Kouni Chahed, M., Ben Salah, A., Bettaieb, J., and Bouafif Ben Alaya, N. (2020). Applying the moving epidemic method to determine influenza epidemic and intensity thresholds using influenza-like illness surveillance data 2009-2018 in Tunisia. Influenza and Other Respiratory Viruses, 14(5):507–514.

Dahlgren, F. S., Shay, D. K., Izurieta, H. S., Forshee, R. A., Wernecke, M., Chillarige, Y., Lu, Y., Kelman, J. A., and Reed, C. (2018). Evaluating oseltamivir prescriptions in centers for Medicare and Medicaid services medical claims records as an indicator of seasonal influenza in the United States. Influenza and Other Respiratory Viruses, 12(4):465–474.

Dahlgren, F. S., Shay, D. K., Izurieta, H. S., Forshee, R. A., Wernecke, M., Chillarige, Y., Lu, Y., Kelman, J. A., and Reed, C. (2019). Patterns of seasonal influenza activity in U.S. core-based statistical areas, described using prescriptions of oseltamivir in Medicare claims data. Epidemics, 26:23 – 31.

Elhakim, M. M., Kandil, S. K., Abd Elaziz, K. M., and Anwar, W. A. (2019). Epidemiology of severe acute respiratory infection (SARI) cases at a sentinel site in Egypt, 2013–15. Journal of Public Health, 42(3):525–533.

Green, H., Charlett, A., Moran-Gilad, J., Fleming, D., Durnall, H., Thomas, D., Cottrell, S., Smyth, B., Kearns, C., Reynolds, A., Smith, G., Elliot, A., Ellis, J., Zambon, M., JM, W., McMenamin, J., and Pebody, R. (2015). Harmonizing influenza primary-care surveillance in the United Kingdom: piloting two methods to assess the timing and intensity of the seasonal epidemic across several general practice-based surveillance schemes. Epidemiology and Infection, 143(1):1–12.

Grilc, E., Prosenc Trilar, K., Lajovic, J., and Sočan, M. (2021). Determining the seasonality of respiratory syncytial virus in Slovenia. Influenza and Other Respiratory Viruses, 15(1):56–63.

Harcourt, S. E., Morbey, R. A., Smith, G. E., Loveridge, P., Green, H. K., Pebody, R., Rutter, J., Yeates, F. A., Stuttard, G., and Elliot, A. J. (2019). Developing influenza and respiratory syncytial virus activity thresholds for syndromic surveillance in England. Epidemiology and Infection, 147:e163.

Lucero, M. G., Inobaya, M. T., Nillos, L. T., Tan, A. G., Arguelles, V. L. F., Dureza, C. J. C., Mercado, E. S., Bautista, A. N., Tallo, V. L., Barrientos, A. V., Rodriguez, T., and Olveda, R. M. (2016). National influenza surveillance in the Philippines from 2006 to 2012: seasonality and circulating strains. BMC Infectious Diseases, 16(1):762.

Ly, S., Arashiro, T., Ieng, V., Tsuyuoka, R., Parry, A., Horwood, P., Heng, S., Hamid, S., Vandemaele, K., Chin, S., Sar, B., and Arima, Y. (2017). Establishing seasonal and alert influenza thresholds in Cambodia using the WHO method: implications for effective utilization of influenza surveillance in the tropics and subtropics. Western Pacific Surveillance and Response Journal, 8:22–32.

Murray, J. L. K., Marques, D. F. P., Cameron, R. L., Potts, A., Bishop, J., von Wissmann, B., William, N., Reynolds, A. J., Robertson, C., and McMenamin, J. (2018). Moving Epidemic Method (MEM) applied to virology data as a novel real time tool to predict peak in seasonal influenza healthcare utilisation. The Scottish experience of the 2017/18 season to date. Eurosurveillance, 23(11):18–00079.

Nisar, N., Aamir, U. B., Badar, N., Mahmood, M. R., Yaqoob, A., Tripathy, J. P., Laxmeshwar, C., Tenzin, K., Zaidi, S. S. Z., Salman, M., and Ikram, A. (2020). Epidemiology of influenza among patients with influenza-like illness and severe acute respiratory illness in Pakistan: A 10-year surveillance study 2008-17. Journal of Medical Virology, 92(12):3028–3037.

Pesälä, S., Virtanen, M. J., Mukka, M., Ylilammi, K., Mustonen, P., Kaila, M., and Helve, O. (2019). Healthcare professionals’ queries on oseltamivir and influenza in Finland 2011-2016 – can we detect influenza epidemics with specific online searches? Influenza and Other Respiratory Viruses, 13(4):364–371.

Páscoa, R., Rodrigues, A. P., Silva, S., Nunes, B., and Martins, C. (2018). Comparison between influenza coded primary care consultations and national influenza incidence obtained by the General Practitioners Sentinel Network in Portugal from 2012 to 2017. PLOS ONE, 13(2):1–10.

Rguig, A., Cherkaoui, I., McCarron, M., Oumzil, H., Triki, S., Elmbarki, H., Bimouhen, A., Falaki, F. E., Regragui, Z., Ihazmad, H., Nejjari, C., and Youbi, M. (2020). Establishing seasonal and alert influenza thresholds in Morocco. BMC Public Health.

Sullivan, S. G., Arriola, C. S., Bocacao, J., Burgos, P., Bustos, P., Carville, K. S., Cheng, A. C., Chilver, M. B., Cohen, C., Deng, Y.-M., El Omeiri, N., Fasce, R. A., Hellferscee, O., Huang, Q. S., Gonzalez, C., Jelley, L., Leung, V. K., Lopez, L., McAnerney, J. M., McNeill, A., Olivares, M. F., Peck, H., Sotomayor, V., Tempia, S., Vergara, N., von Gottberg, A., Walaza, S., and Wood, T. (2019). Heterogeneity in influenza seasonality and vaccine effectiveness in Australia, Chile, New Zealand and South Africa: early estimates of the 2019 influenza season. Eurosurveillance, 24(45).

Tay, E. L., Grant, K., Kirk, M., Mounts, A., and Kelly, H. (2013). Exploring a proposed WHO method to determine thresholds for seasonal influenza surveillance. PLOS ONE, 8(10).

Torner, N., Basile, L., Martínez, A., Rius, C., Godoy, P., Jané, M., Domínguez, A., Aizpurua, J., Alonso, J., Azemar, J., Aizpurua, P., Ardaya, P. M., Basas, M. D., Batalla, J., Biendicho, P., Bonet, M., Callado, M., Campos, S., Casanovas, J. M., Ciurana, E., Clapes, M., Cots, J. M., De la Rica, D., Domingo, I., Elizalde, G., Escapa, P., Fajardo, S., Fau, E., Fernandez, O., Fernandez, M., Ferrer, C., Forcada, A., Fos, E., Gadea, G., Garcia, J., Garcia, R., Gatius, C., Gelado, M. J., Grau, M., Grivé, M., Guzman, M. C., Hernández, R., Jimenez, G., Juscafresa, A., LLussa, A. M., López, C., Kristensen, L., Macià, E., Mainou, A., Marco, E., Martínez, M., Martínez, J. G., Marulanda, K. V., Masa, R., Moncosí, X., Naranjo, M. A., Navarro, D., Ortolà, E., París, F., Pérez, M. M., Pozo, C., Pujol, R., Ribatallada, A., Ruiz, G., Sabaté, S., Sanchez, R., Sarrà, N., Tarragó, E., Teixidó, A. M., Torres, A., Valén, E., Van Esso, D., Van Tarjcwick, C., Vink Schoenholzer, R., Zabala, E., Marcos, M. A., Mosquera, M. D. M., de Molina, P., Rubio, E., Isanta, R., Anton, A., Pumarola, T., Vilella, A., Gorrindo, P., Espejo, E., Andrés, M., Barcenilla, F., Navarro, G., Barrabeig, I., Pou, J., Alvarez, P., Plasencia, E., Rebull, J., Sala, M. R., Riera, M., Camps, N., Follia, N., Oller, A., Godoy, P., Bach, P., Rius, C., Hernández, R., Perez, R., Torra, R., Carol, M., Minguell, S., Marce, R., Garcia-Pardo, G., Olona, M., Alvarez, A., Ramon, J. M., Mòdol, J. M., Mena, G., Campins, M., Massuet, C., Tora, G., Ferràs, J., Ferrús, G., and The Working Group on PIDIRAC Sentinel Surveillance of Catalonia (2019). Assessment of two complementary influenza surveillance systems: sentinel primary care influenza-like illness versus severe hospitalized laboratory-confirmed influenza using the moving epidemic method. BMC Public Health, 19(1089).

Vette, K., Bareja, C., Clark, R., and Lal, A. (2018). Establishing thresholds and parameters for pandemic influenza severity assessment, Australia. Bulletin of the World Health Organization, 96:558–567.

Wagner, M., Lampos, V., Cox, I. J., and Pebody, R. (2018). The added value of online user-generated content in traditional methods for influenza surveillance. Scientific Reports, 8(1):13963.

